# Specifications of the ACMG/AMP variant interpretation guidelines for germline *TP53* variants

**DOI:** 10.1101/2020.04.25.20078931

**Authors:** Cristina Fortuno, Kristy Lee, Magali Olivier, Tina Pesaran, Phuong L. Mai, Kelvin C. de Andrade, Laura D. Attardi, Stephanie Crowley, D. Gareth Evans, Bing-Jian Feng, Ann Katherine Major Foreman, Megan N. Frone, Robert Huether, Paul A. James, Kelly McGoldrick, Jessica Mester, Bryce A. Seifert, Thomas P. Slavin, Leora Witkowski, Liying Zhang, Sharon E. Plon, Amanda B. Spurdle, Sharon A. Savage, on behalf of the ClinGen *TP53* Variant Curation Expert Panel

## Abstract

Germline pathogenic variants in *TP53* are associated with Li-Fraumeni syndrome (LFS), an autosomal dominant cancer predisposition disorder associated with high risk of malignancy, including early onset breast cancers, sarcomas, adrenocortical carcinomas and brain tumors. Intense cancer surveillance for individuals with *TP53* germline pathogenic variants has been shown to decrease mortality; therefore, accurate and consistent classification of variants across clinical and research laboratories is crucial to patient care. Here, we describe the work performed by the Clinical Genome Resource *TP53* Variant Curation Expert Panel (ClinGen *TP53* VCEP) focused on specifying the American College of Medical Genetics and Genomics and the Association for Molecular Pathology (ACMG/AMP) guidelines for germline variant classification to the *TP53* gene. Specifications were applied to twenty ACMG/AMP criteria while nine were deemed not applicable. The original strength level for ten criteria was also adjusted due to current evidence. Use of the *TP53*-specific guidelines and sharing of clinical data amongst experts and clinical laboratories led to a decrease in variants of uncertain significance from 28% to 12% in comparison with the original guidelines. The ClinGen *TP53* VCEP recommends the use of these *TP53*-specific ACMG/AMP guidelines as the standard strategy for *TP53* germline variant classification.

## INTRODUCTION

The *TP53* gene encodes p53, a protein with essential roles in genome stability and key cellular functions such as cell cycle, metabolism, apoptosis, senescence and differentiation (Lane, 1992; Zerdoumi et al., 2017). Germline pathogenic variants in *TP53* occur in ∼70% of individuals with Li-Fraumeni syndrome (LFS) (Schneider, Zelley, Nichols, & Garber, 1993), defined by patterns of personal and family history of certain early onset cancers, mainly pre-menopausal breast cancer, bone and soft tissue sarcomas, adrenocortical carcinomas and brain tumors. The cumulative incidence of cancer in LFS families is nearly 50% by age 31y for females and 46y for males, and up to 100% by age 70y for both (Mai et al., 2016). The extremely high penetrance has led to recommendations for intensive surveillance and other clinical management strategies (Ballinger et al., 2017; Evans, Birch, Ramsden, Sharif, & Baser, 2006; Kratz et al., 2017; Schon & Tischkowitz, 2018; Villani et al., 2016). Reported prevalence estimates range from 1:5,000 to 1:20,000 (Gonzalez, Noltner, et al., 2009; Lalloo et al., 2006), but recent studies suggest the frequency of germline pathogenic *TP53* variants may be higher (Amendola et al., 2015; de Andrade et al., 2019; de Andrade et al., 2017).

Germline *TP53* genetic testing was initially recommended for individuals meeting Classic LFS (Li et al., 1988) and/or Chompret criteria, most recently revised in 2015 criteria (Bougeard et al., 2015). Advances in next-generation sequencing have expanded the use of multigene panels as well as the inclusion of *TP53* as a secondary finding for genome-scale sequencing (Kalia et al., 2017). This has led to an increased number of variants of uncertain significance identified in *TP53*, including in cancer patients who do not meet LFS criteria, and individuals without cancer (Bittar et al., 2019). Given the significant clinical and emotional challenges that come with an LFS diagnosis (Ballinger et al., 2017; Evans et al., 2006; Kratz et al., 2017; Schon & Tischkowitz, 2018; Villani et al., 2016; Young et al., 2019), it is essential to correctly assign *TP53* germline variant pathogenicity.

The American College of Medical Genetics and Genomics and the Association for Molecular Pathology (ACMG/AMP) variant curation guidelines are a series of generic criteria with different levels of strength for and against pathogenicity, incorporating evidence from multiple data sources(Richards et al., 2015). As part of the directive of the Clinical Genome consortium (ClinGen: https://clinicalgenome.org), specifications of these guidelines for specific gene/disease pairs are developed and documented by a Variant Curation Expert Panel (VCEP), and have been completed for cancer genes including *CDH1* (Lee et al., 2018) and *PTEN* (Mester et al., 2018).

Herein, we present the scientific rationale and recommendations of the ClinGen *TP53* VCEP to adapt the ACMG/AMP guidelines for the classification of *TP53* germline variants and present results from pilot testing of the finalized guidelines.

## METHODS

The *TP53* VCEP followed ClinGen standard operating procedures (see https://clinicalgenome.org/curation-activities/variant-pathogenicity/documents/). The VCEP formed in 2015 by recruiting international *TP53* experts knowledgeable in phenotype, molecular etiology, and functional processes. Twenty-four members contributed to at least one of three different evidence type working groups: Population/Computational, Functional, and Clinical. Each group reviewed the ACMG/AMP specifications in detail, incorporated this with critical review of the relevant literature and analysis of relevant data to inform evidence weights, and came to consensus for each specification.

VCEP members nominated variants for pilot testing the *TP53*-specific ACMG/AMP guidelines; 23 variants were selected to cover different molecular effects and the availability of data to allow assessing usability of different criteria. Seven variants from the International Agency on Research in Cancer (IARC) *TP53* Database (Bouaoun et al., 2016) were included for their rich phenotypic information, but had no prior variant classifications. Thirteen variants from ClinVar database (Landrum et al., 2018) were used to balance the spectrum of suspected benign to pathogenic variants. Case level evidence available from clinical, research or clinical laboratory databases were provided, including information regarding cancer type(s) and family history, familial variant segregation, and *de novo* observations. Variant classifications (pathogenic (P), likely pathogenic (LP), variant of uncertain significance (VUS), likely benign (LB) and benign (B)) were also provided, which are referred to as prior expert assertions. Of note, variants pulled from ClinVar had assertions submitted by laboratories with representation on our VCEP, so that laboratory’s assertion in 2017 was considered as the prior expert assertion. Each variant was independently curated by two the five biocurators using the original ACMG/AMP guidelines and the *TP53* specifications to test user interpretability. The criteria combinations for a given classification tier were as originally proposed (Richards et al., 2015), with the additional combination of very strong plus supporting criterion reaching LP for the *TP53*-specific guidelines supported by the Tavtigian et al. Bayesian rule combination calculator (Tavtigian et al., 2018). The resulting classifications were compared between biocurators, against prior assertions by the nominating expert(s) or contributing laboratories, and with ClinVar assertions (Landrum et al., 2018) when possible. During this phase, the *TP53*-specific guidelines were refined, and the final draft was shared with the ClinGen Sequence Variant Interpretation (SVI) Committee for approval.

## RESULTS

### *TP53*-specific variant curation criteria

Final *TP53-*specific ACMG/AMP guidelines are summarized in Table 1. Of the 28 original criteria, nine (PM3, PM4, PP2, PP4, PP5, BP1, BP3, BP5, and BP6) were excluded due to either limited data to support use of a rule code, irrelevance to *TP53*, or to avoid redundancy with another criterion specification. Additional details are shown in Supplementary Table S1. The remaining 19 criteria were modified by detailing the content and/or changing the strength level. Rationale for criteria specification are explained below.

**Table 1.** **Summary of the *TP53*-specificACMG/AMP criteria developed by the ClinGen *TP53* Variant Curation Expert Panel**

| Original ACMG/AMP guidelines |  | *TP53* specifications |
| --- | --- | --- |
| Rule code | Criteria description |  |
| **PVS1** | Null variant (nonsense, frameshift, canonical $\pm 1$ or 2 splice sites, initiation codon, single or multiexon deletion) in a gene where LOF is a known mechanism of disease | Use SVI-approved decision tree to determine the strength of this criterion (refer to Abou Tayoun et al. for more details). |
| **PS1** | Same amino acid change as a previously established pathogenic variant regardless of nucleotide change | Use original description with the following additions:<br>**PS1:**<br>- Must confirm there is no difference in splicing using RNA data.<br>- Can only be used to compare to variants classified as Pathogenic or Likely Pathogenic by the *TP53* VCEP (see ClinVar for VCEP classifications).<br>**PS1_Moderate:**<br>- Must confirm there is no difference in splicing using *in silico* modelling data (MaxEntScan and Human Splicing Finder).<br>- Can only be used to compare to variants classified as Pathogenic or Likely Pathogenic by the *TP53* VCEP (see ClinVar). |
| **PS2** | *De novo* (both maternity and paternity confirmed) in a patient with the disease and no family history | Use SVI-approved scoring system to determine the strength of this criterion (refer to Table 7 for more details) |
| **PS3** | Well-established *in vitro* or *in vivo* functional studies supportive of a damaging effect on the gene or gene product | **PS3:** transactivation assays in yeast demonstrate a low functioning allele ($\leq 20\%$ activity) **AND** there is evidence of dominant negative effect and loss-of-function **OR** there is a second assay showing low function (colony formation assays, apoptosis assays, tetramer assays, knock-in mouse models and growth suppression assays).<br>**PS3_Moderate:** transactivation assays in yeast demonstrate a partially functioning allele ($>20\%$ and $\leq 75\%$ activity) **AND** there is evidence of dominant negative effect and loss-of-function **OR** there is a second assay showing low function (colony formation assays, apoptosis assays, tetramer assays, knock-in mouse models and growth suppression assays). |
|  |  | **PS3_Moderate:** there is no data available from transactivation assays in yeast **BUT** there is evidence of dominant negative effect and loss-of-function **AND** there is a second assay showing low function (colony formation assays, apoptosis assays, tetramer assays, knock-in mouse models and growth suppression assays).<br>Refer to Figure 1 for more details. |
| **PS4** | The prevalence of the variant in affected individuals is significantly increased compared with the prevalence in controls | Use SVI-approved scoring system to determine the strength of this criterion (refer to Table 3 for more details). This criterion cannot be applied when a variant also meets BA1 or BS1. Refrain from considering probands who have another pathogenic variant(s) in a highly penetrant cancer gene(s) that is a logical cause for presentation.<br><u>Caveat:</u> Please be mindful of the risk of clonal hematopoieses of indeterminate potential with *TP53* variants (Coffee et al., 2017; Weitzel et al., 2017). One should take care to ensure that probands have germline and not mosaic somatic *TP53* variants. |
| **PM1** | Located in a mutational hot spot and/or critical and well-established functional domain (e.g., active site of an enzyme) without benign variation | Located in a mutational hotspot defined as:<br>- Variants within the following codons on protein NP_000537.3: 175, 273, 245, 248, 282, 249.<br>- Variants seen in cancerhotspots.org (v2) with >10 somatic occurrences (recommendation from the ClinGen Germline/Somatic Variant Curation Subcommittee). |
| **PM2** | Absent from controls (or at extremely low frequency if recessive) in Exome Sequencing Project, 1000 Genomes Project, or Exome Aggregation Consortium | **PM2_Supporting:** absent from population databases (gnomAD (most up-to-date non-cancer dataset) is the preferred population database at this time http://gnomad.broadinstitute.org). |
| **PM3** | For recessive disorders, detected *in trans* with a pathogenic variant | Excluded (refer to Supplementary Table 1 for more details). |
| **PM4** | Protein length changes as a result of in-frame deletions/insertions in a nonrepeat region or stop-loss variants | Excluded (refer to Supplementary Table 1 for more details). |
| **PM5** | Novel missense change at an amino acid residue where a different missense change determined to be pathogenic has been seen before | **PM5:** novel missense change at an amino acid residue where at least two other different missense changes determined to be pathogenic by the *TP53* VCEP have been seen before.<br>**PM5_Supporting:** novel missense change at an amino acid residue where a different missense change determined to be pathogenic by the *TP53* VCEP has been seen before.<br>Both criteria require the following additions:<br>- Grantham should be used to compare the variants, and the variant being evaluated must have equal to or higher score than the observed pathogenic variants.<br>- Splicing should be ruled out using *in silico* modelling data (MaxEntScan and Human Splicing Finder).<br>- This criterion cannot be applied when a variant also meets PM1. |
| **PM6** | Assumed *de novo*, but without confirmation of paternity and maternity | Use SVI-approved scoring system to determine the strength of this criterion (refer to Table 7 for more details). |
| **PP1** | Co-segregation with disease in multiple affected family members in a gene definitively known to cause the disease | PP1: co-segregation with disease is observed in 3-4 meioses in one family.<br>PP1_Moderate: co-segregation with disease is observed in 5-6 meioses in one family.<br>PP1_Strong: co-segregation with disease is observed >7 meioses in >1 family. |
| **PP2** | Missense variant in a gene that has a low rate of benign missense variation and in which missense variants are a common mechanism of disease | Excluded (refer to Supplementary Table 1 for more details). |
| **PP3** | Multiple lines of computational evidence support a deleterious effect on the gene or gene product (conservation, evolutionary, splicing impact, etc.) | PP3: Use original description with the following additions:<br>- For missense variants, use a combination of BayesDel ($\geq 0.16$) and optimised Align-GVGD (C55-C25).<br>- For splicing variants, use MaxEntScan and Human Splicing Finder.<br>PP3_Moderate: for missense variants, use a combination of BayesDel ($\geq 0.16$) and optimized Align-GVGD (C65). |
| **PP4** | Patient's phenotype or family history is highly specific for a disease with a single genetic etiology | Excluded (refer to Supplementary Table 1 for more details). |
| **PP5** | Reputable source recently reports variant as pathogenic, but the evidence is not available to the laboratory to perform an independent evaluation | Excluded (refer to Supplementary Table 1 for more details). |
| **BA1** | Allele frequency is >5% in Exome Sequencing Project, 1000 Genomes Project, or Exome Aggregation Consortium | Allele frequency is $\geq 0.1\%$ in a non-founder population with a minimum of five alleles (gnomAD (most up-to-date non-cancer dataset)) is the preferred population database at this time<br>http://gnomad.broadinstitute.org). |
| **BS1** | Allele frequency is greater than expected for disorder | Allele frequency is $\geq 0.03\%$ and $< 0.1\%$ in a non-founder population with a minimum of five alleles (gnomAD (most up-to-date non-cancer dataset) is the preferred population database at this time<br>http://gnomad.broadinstitute.org). |
| **BS2** | Observed in a healthy adult individual for a recessive (homozygous), dominant (heterozygous), or X-linked (hemizygous) disorder, with full penetrance expected at an early age | BS2: observed in a single dataset in $\geq 8$ females, who have reached at least 60 years of age without cancer (i.e. cancer diagnoses after age 60 are ignored).<br>BS2_Supporting: observed in a single dataset in 2-7 females, who have reached at least 60 years of age without cancer. |
|  |  | Caveat: Be mindful of the risk of clonal hematopoiesis of indeterminate potential with *TP53* variants (Coffee et al., 2017; Weitzel et al., 2017). Individuals with mosaic somatic *TP53* variants should not be included as evidence for BS2. |
| **BS3** | Well-established *in vitro* or *in vivo* functional studies show no damaging effect on protein function or splicing | - BS3: transactivation assays in yeast demonstrate a functional allele or super-transactivation (>75% activity) **AND** there is no evidence of dominant negative effect and loss-of-function **OR** there is a second assay showing retained function (colony formation assays, apoptosis assays, tetramer assays, knock-in mouse models and growth suppression assays).<br>- BS3_Supporting: transactivation assays in yeast demonstrate a partially functioning allele (>20% and ≤75% activity) **AND** there is no evidence of dominant negative effect and loss-of-function **OR** there is a second assay showing retained function (colony formation assays, apoptosis assays, tetramer assays, knock-in mouse models and growth suppression assays).<br>- BS3_Supporting: there is no data available from transactivation assays in yeast **BUT** there is no evidence of dominant negative effect and loss-of-function **AND** there is a second assay showing retained function (colony formation assays, apoptosis assays, tetramer assays, knock-in mouse models and growth suppression assays).<br>Refer to Figure 3 for more details. |
| **BS4** | Lack of segregation in affected members of a family | The variant segregates to opposite side of the family meeting LFS criteria, or the variant is present in >3 living unaffected individuals (at least two of three should be female) above 55 years of age. |
| **BP1** | Missense variant in a gene for which primarily truncating variants are known to cause disease | Excluded (refer to Supplementary Table 1 for more details). |
| **BP2** | Observed *in trans* with a pathogenic variant for a fully penetrant dominant gene/disorder or observed *in cis* with a pathogenic variant in any inheritance pattern | Variant is observed *in trans* with a *TP53* pathogenic variant (phase confirmed), or there are three or more observations with a *TP53* pathogenic variant when phase is unknown (at least two different *TP53* pathogenic variants). The other observed pathogenic variants must have been classified using the *TP53*-specific guidelines. |
| **BP3** | In-frame deletions/insertions in a repetitive region without a known function | Excluded (refer to Supplementary Table 1 for more details). |
| **BP4** | Multiple lines of computational evidence suggest no impact on gene or gene product (conservation, evolutionary, splicing impact, etc.) | Same rule description with the following additions:<br>- For missense variants, use a combination of BayesDel (<0.16) and optimized Align-GVGD (C15-C0).<br>- For splicing variants, use MaxEntScan and Human Splicing Finder. |
| **BP5** | Variant found in a case with an alternate molecular basis for disease | Excluded (refer to Supplementary Table 1 for more details). |
| **BP6** | Reputable source recently reports variant as benign, but the evidence is not available to the laboratory to perform an independent evaluation | Excluded (refer to Supplementary Table 1 for more details). |
| **BP7** | A synonymous (silent) variant for which splicing prediction algorithms predict no impact to the splice consensus sequence nor the creation of a new splice site AND the nucleotide is not highly conserved | Same description with the following additions:<br>- Splicing should be ruled out using *in silico* modelling data (MaxEntScan and Human Splicing Finder).<br>- If a new alternate site is predicted, compare strength to native site in interpretation. |

Abbreviations: SVI = Sequence Variant Interpretation; VCEP = Variant Curation Expert Panel

### Population/Computational Working Group

#### Population data

BS1 and BA1 are criteria against pathogenicity based on the frequency of a germline variant in healthy individuals. To define the *TP53* variant frequency cutoff for BS1, we calculated the maximum credible population allele frequency as reported by Whiffin et al. (Whiffin et al., 2017). Conservative values were used for prevalence and penetrance of *TP53*, which are 1 in 5,000 individuals (Lalloo et al., 2006) and 30% cancer risk similarly used by other hereditary cancer VCEPs (Lee et al., 2018; Mester et al., 2018). Genetic heterogeneity was set at 1 as *TP53* is the only LFS-associated gene. This resulted in BS1 at 0.03%. BA1 was then defined as 0.1% based on a less conservative lifetime penetrance estimate of 70%, holding allelic and genetic heterogeneity at 1, and then increasing the derived maximum credible population allele frequency of 0.01% by on order of magnitude to arrive at a cutoff of 0.1%. For both criteria, a minimum of five alleles in a given population was required to ensure a comparable cohort. The *TP53* VCEP specifically recommends the use of the most up-to-date non-cancer dataset of the gnomAD database (Karczewski et al., 2019), as this is the largest control database currently publicly available, and to ignore frequencies in Ashkenazi Jewish and Finnish populations due to founder effects (Kaariainen, Muilu, Perola, & Kristiansson, 2017; Shi et al., 2017).

The PM2 criterion uses absence in controls as evidence towards pathogenicity. Due to the overall rarity of *TP53* germline variants (benign or pathogenic), the *TP53* VCEP downgraded this criterion to supporting strength level.

PS4 requires case-control analyses, considered impractical for *TP53* due to variant rarity and limited number of published studies. We instead developed a proband counting system to assign pathogenicity, based on the number of variant carriers meeting each of the existing clinical criteria for LFS. A likelihood ratio (LR) towards pathogenicity was calculated by dividing the proportion of carriers meeting classic LFS or Chompret 2015 criteria by the proportion of non-carriers meeting the same criteria, using data from individuals undergoing multigene panel testing at Ambry Genetics (Supplementary Figure S1). Results indicated that one proband with a variant meeting classic LFS or Chompret 2015 criteria would provide enough evidence to reach moderate (LR=15.57) or supporting (LR=3.37) strength level, respectively (Tavtigian et al., 2018). However, given the width of the confidence intervals, more conservative weights were assigned using a point system based on the number of probands meeting classic LFS or Chompret 2015 (Table 2). Additionally, it was decided that PS4 should not be applied if the variant also meets the population rules BS1 or BA1, to avoid coincidental accumulation of proband counts for common variants, following the approach previously developed for *PTEN* (Mester et al., 2018).

**Table 2.** **Point system created for the specifications of the PS4 rule\***

| **Classic LFS** | **PS4 Evidence Strength** | **# of Points Required** |
| --- | --- | --- |
| 1 point for each proband | PS4 | 4 or more points |
| **Chompret 2015** | PS4_Moderate | 2-3 points |
| 0.5 point for each proband | PS4_Supporting | 1 point |

\*Not to be used if a variant also meets BS1 or BA1 rules.

The BS2 criterion uses presence of a variant in unaffected adults as evidence against pathogenicity. Two independent datasets (Ambry Genetics and GeneDx) were used to calculate how many observations of cancer-free adults at age 60 equated to strong evidence against pathogenicity. The LR towards pathogenicity was calculated by comparing the proportion of these individuals with a known *TP53* pathogenic variant versus individuals without a *TP53* pathogenic variant (Supplementary Table S2). The LRs estimated for observation of a variant in one healthy individual were 0.66 (Ambry dataset) and 0.28 (GeneDx dataset). Selecting the more conservative LR of 0.66, two to seven healthy individuals were considered necessary to apply BS2_Supporting (LRs ranging from 0.44 to 0.06), and eight or more to apply BS2 (LRs lower than 0.04) (Tavtigian et al., 2018). Given that the dataset used for this analysis included mostly female, and the associated risk of pre-menopausal breast cancer (Mai et al., 2016) for female *TP53* carriers elevates their cancer risk compared with male *TP53* carriers, it was stipulated that this criterion should be applied to female carriers only.

#### Computational and predictive data

PP3 and BP4 are commonly used criteria related to predictions using bioinformatic tools. Based on previous findings (Fortuno, James, Young, et al., 2018), an optimized version of Align-GVGD (Mathe et al., 2006) combined with BayesDel (Feng, 2017) (both included in the IARC *TP53* Database R20 (Bouaoun et al., 2016)) were selected for bioinformatic prediction of *TP53* missense variant pathogenicity. To assess prediction of effects on splicing, we selected MaxEntScan (Yeo & Burge, 2004) and Human Splicing Finder (HSF) (Desmet et al., 2009), commonly used and freely available tools that have shown good performance for predicting variant spliceogenicity for other cancer syndrome genes (Shamsani et al., 2019).

The BP7 criterion for silent variants was expanded to specify the use of MaxEntScan and HSF to predict any effects on splicing.

Strength level for PM5, which relies on previous observations of pathogenic variants at a given location as evidence of pathogenicity for other variants at the same location, was assessed as follows: The 493 non-functional variants from yeast transactivation assays(Kato et al., 2003) were assessed for the number of additional variants at the same amino acid residue that were also reported as non-functional, and compared to the number of functional or super trans variants (N=1239) at that position to generate a LR towards pathogenicity (Supplementary Table S3). Results indicated PM5 was applicable if two other pathogenic variants have been seen at a given amino acid residue (LR=6.46), but should be downgraded to supporting strength level if only one other pathogenic variant has been seen at the same residue (LR=2.90) (Tavtigian et al., 2018). Further, the following restrictions were added: (i) known P/LP variants must be based on classifications using the *TP53*-specific guidelines; (ii) variants using this rule must have equal or higher Grantham score (Grantham, 1974) than at least one pathogenic variant observed at that codon; (iii) this criterion cannot be used for any variant for which PM1 has been applied; (iv) splicing effects are excluded based on bioinformatic evidence from MaxEntScan and HSF.

PS1 may be applied as strong strength if effects on splicing due to the nucleotide change are excluded using splicing assay data, or downgraded to PS1_Moderate if only bioinformatic predictions (using MaxEntScan and HSF) are available as evidence against aberrant splicing. The comparative variant must have been classified as P/LP using the *TP53*-specific guidelines.

PVS1 is the only original criterion with a very strong strength level for pathogenicity, which applies to loss of function variants. The ClinGen SVI has published further guidance on this rule (Abou Tayoun et al., 2018), for which the *TP53* VCEP has agreed to follow, including applying relative strengths for different types of null variants based on characteristics, such as variant type and location.

### Functional Working Group

#### Functional data

The functional-related PS3 and BS3 criteria, as originally described, do not specify required functional assays for a given gene. There are a number of published systematic functional studies of p53 missense variants, which measure transactivation activity(Kato et al., 2003), loss of function (LOF) (Giacomelli et al., 2018; Kotler et al., 2018) and dominant-negative effect (DNE) (Giacomelli et al., 2018). Matthews correlation coefficient was used to determine the relative performance of the different assay results for variant pathogenicity prediction; clinical reference sets were assumed pathogenic missense variants (present in classic LFS probands in the IARC *TP53* Database R19 and absent in controls, N=52) and assumed benign missense variants (in controls from non-cancer gnomAD v2.1.1 or FLOSSIES (https://whi.color.com/) at a frequency higher than 0.0001 and not found in patients, N=31) (Supplementary Table 4). A decision tree considering availability and relative performance of the different assays is shown in Figure 3.

#### Hotspot data

The second part of the PM1 criterion related to protein regions was considered not applicable, as there is no known functional domain without benign variation in *TP53*. However, there are several well-described hotspots in *TP53*, occurring at amino acid positions 175, 245, 248, 249, 273, 282 (Brosh & Rotter, 2009; Fortuno, Pesaran, et al., 2019; Freed-Pastor & Prives, 2012), for which PM1 is applicable. Additionally, analysis of tumor DNA sequencing has identified a large number of *TP53* variants as somatic hotspots, with information available at cancerhotspots.org (Chang et al., 2016; Chang et al., 2018). Following recommendations from the ClinGen Germline/Somatic Variant Curation Subcommittee (Walsh et al., 2018), it was specified that the PM1 criterion can also apply to somatically detected hotspots with ≥10 occurrences in cancerhotspots.org. This information is annotated in the IARC *TP53* Database R20(Bouaoun et al., 2016).

### Clinical Working Group

#### Segregation data

The original ACMG/AMP criteria use segregation as supporting strength for pathogenicity (PP1), allowing for stronger evidence if there is more segregation data, or a strong strength of evidence against pathogenicity when there is lack of segregation (BS4). Given the wide spectrum of cancer types that have been reported for *TP53* carriers (Caron et al., 2016; Kratz et al., 2017; Olivier, Hollstein, & Hainaut, 2010), the criteria were specified based only on the number of meiosis of any LFS-associated cancer type, and number of families reported. Based on the gradations created previously for *PTEN*, the resulting *TP53* specifications for PP1 are detailed in Table 1.

For BS4, which uses lack of segregation as evidence against pathogenicity, we considered potential issues due to the high *de novo* rate reported for *TP53*(Gonzalez, Buzin, et al., 2009), and specified use under these two scenarios: the variant segregates to the side of the family that does not meet LFS criteria, or the variant is present in three or more living unaffected individuals (where at least two of three are female) above 55 years of age (age specification consistent with Chompret 2015 criteria).

#### De novo data

The *TP53* VCEP provides guidance for assigning the strength for the original *de novo* PS2 and PM6 criteria that should be based on the type of cancer and its relevance to the *TP53* spectrum. This was accomplished using a point system incorporating parentage and proband cancer type which also allows for combination of points if there are multiple *de novo* reports of the same variant (Table 3).

**Table 3.** **Point systems created for the specifications of the PS2 and PM6 criteria and *TP53*-associated cancers with different strength levels\***

|  |  |  |  |
| --- | --- | --- | --- |
| **Strong LFS criteria**<br>• 2 points for each cancer in *de novo* individual when maternity and paternity are confirmed<br>• 1 point for each cancer when parental testing is not available |  | • Breast cancer (IDC & DCIS) <31 years of age<br>• Choroid plexus carcinoma<br>• Adrenocortical adenoma or carcinoma <18 years of age<br>• Rhabdomyosarcoma <46 years of age<br>• Osteosarcoma <46 years of age |  |
| **Moderate LFS criteria**<br>• 1 point for each cancer in *de novo* individual when maternity and paternity are confirmed<br>• 0.5 point for each cancer when parental testing is not available |  | • Breast cancer >30 and <50 years of age<br>• Malignant brain tumors (excluding optic gliomas) <46 years of age<br>• Primary lung cancer <46 years of age<br>• Adrenocortical adenoma or carcinoma ≥18 and <50 years of age<br>• Rhabdomyosarcoma or osteosarcoma >45 years of age<br>• Other sarcomas (*e.g.*, malignant phyllodes tumor, leiomyosarcoma, liposarcoma) <60 years of age<br>○ Exclude dermatofibrosarcoma & Ewing sarcoma<br>• Hypodiploid ALL (Specifically low-hypodiploid 32-39 chromosomes) |  |
| Total points required to assign the following rule codes |  |  |  |
| **PM6_Supporting** | **PS2_Moderate or PM6** | **PS2 or PM6_Strong** | **PS2_VeryStrong or PM6_VeryStrong** |
| 0.5 | 1 | 2 | 4 |

\*If there are multiple reports of *de novo* probands, the points for each *de novo* observation can be summed. If the proband has multiple cancers, only the strongest associated LFS cancer should be used.

**Table 4.** Variant classified during the pilot testing phase using the *TP53*-specific ACMG/AMP guidelines. All variants were annotated in relation to the transcript NM_000546.5 and protein NP_000537.3.

| *TP53* variant | ClinVar ID | ClinVar Classifications (September, 2019)* | Prior Variant Classifications^ | Dual Biocurator Classifications |  | *TP53*-specific ACMG/AMP Final Classifications | Final Criteria Applied |
| --- | --- | --- | --- | --- | --- | --- | --- |
|  |  |  |  | Original ACMG/AMP Guidelines# | *TP53*-specific ACMG/AMP Guidelines% |  |  |
| c.1079G>C<br>p.(Gly360Ala) | 142003 | B/LB | LB | B/VUS | LB | LB | BS1, BS3_Supporting, BP4 |
| c.883C>T<br>p.(Pro295Ser) | 428862 | LB | LB | VUS | LB | VUS | BP4 |
| c.206C>G<br>p.(Ala69Gly) | 230112 | LB | LB | VUS | VUS | VUS | PM2_Supporting, BP4 |
| c.1093C>T<br>p.(His365Tyr) | 80708 | VUS | VUS | VUS | LB | LB | BS3, BP4 |
| c.403T>G<br>p.(Cys135Gly) | 376563 | VUS | VUS | LP | VUS | LP | PS3, PM2_Supporting, PP3_Moderate |
| c.1000G>C<br>p.(Gly334Arg) | 182969 | VUS | VUS | VUS | VUS | VUS | BS3, PP3, PS4_Supporting |
| c.532C>G<br>p.(His178Asp) | 482223 | LP | NA | LP/P | LP/P | LP | PS3, PM2_supporting, PM6, PP3_Moderate |
| c.538G>A<br>p.(Glu180Lys) | 245711 | LP | P | P | VUS | LP | PM2_Supporting, PP3, PS4_Supporting, PM6, PS3_Moderate |
| c.431A>T<br>p.(Gln144Leu) | 376647 | LP | NA | VUS | VUS | VUS | PM2_Supporting, PS4_Supporting, BS3_Supporting |
| c.578A>C<br>p.(His193Pro) | 376612 | LP | NA | P | P | P | PS3, PM6, PM2_Supporting, PP3_Moderate, PS4_Supporting |
| c.743G>T<br>p.(Arg248Leu) | 230253 | LP | P | P/LP | LP | P | PS3, PM1, PM2_Supporting, PP3_Moderate, PS4_Supporting |
| c.97-1G>A | 638853 | LP | P | P | LP/P | P | PVS1_Strong, PM2_Supporting, PS4_Supporting |
| c.537T>A<br>p.(His179Gln) | 406578 | P/LP | NA | P | P | LP | PM6, PS3, PM2_Supporting, PP3 |
| c.517G>A<br>p.(Val173Met) | 233951 | P/LP | LP | P | LP/P | P | PS2, PS3, PM2_Supporting, PS4_Supporting, PP3 |
| c.743G>A<br>p.(Arg248Gln) | 12356 | P/LP | P | P | LP | P | PS3, PM1, PP3, PS4, PS2 |
| c.818G>A<br>p.(Arg273His) | 12366 | P/LP | P | P | P | P | PS3, PS2, PS4, PM1, PP3 |
| c.1031T>C<br>p.(Leu344Pro) | 12375 | P | LP | P/LP | LP | P | PS3,<br>PM2_Supporting,<br>PP3_Moderate,<br>PS4_Moderate |
| c.659A>G<br>p.(Tyr220Cys) | 127819 | P | P | P | P | P | PS3, PP1_strong,<br>PM6,<br>PP3_Moderate,<br>PS4_Moderate |
| c.742C>T<br>p.(Arg248Trp) | 12347 | P | P | P | P | P | PS2, PS3,<br>PP1_Strong,<br>PM1, PS4,<br>PP3_Moderate |
| c.993+1delG | 428898 | P | P | P | P | P | PVS1,<br>PM2_Supporting,<br>PS4_Supporting,<br>PP1_Moderate |
| c.892G>T<br>p.(Glu298*) | 93323 | P | P | P | P | P | PVS1,<br>PM2_Supporting,<br>PS4_Supporting |
| c.372C>A<br>p.(Cys124*) | 458537 | P | NA | P | P | P | PVS1, PM6,<br>PM2_Supporting |
| c.488A>G<br>p.(Tyr163Cys) | 127814 | P | NA | LP/P | P | P | PS3,<br>PS2_Moderate,<br>PM2_Supporting,<br>PP3_Moderate,<br>PS4_Supporting |
| c.455C>T<br>p.(Pro152Leu) | 142766 | P | P | P | LP/P | P | PS3,<br>PP3_Moderate,<br>PS4, PP1 |
| c.919+1G>A | 633606 | P | P | P | P | P | PVS1_Strong,<br>PM6_Supporting,<br>PM2_Supporting |
| c.733G>A<br>p.(Gly245Ser) | 12365 | P | P | P | P | P | PS3, PM1, PP3,<br>PS4 |
| c.869G>A<br>p.(Arg290His) | 127825 | Conflicting | VUS | VUS | VUS | B | BS3, BP4, BS1 |
| c.847C>T<br>p.(Arg283Cys) | 127824 | Conflicting | VUS | VUS | VUS | VUS | BS3, PP3 |
| c.1040C>A<br>p.(Ala347Asp) | 43587 | Conflicting | P | P | LP/P | LP | PS3,<br>PM2_Supporting,<br>PP1_Moderate,<br>PS4_Supporting |
| c.1120G>C<br>p.(Gly374Arg) | 230269 | Conflicting | LB/VUS | LB/VUS | LB | LB | BS3, BP4 |
| c.1096T>G<br>p.(Ser366Ala) | 135360 | Conflicting | LB/VUS | LB/VUS | LB | LB | BS3, BP4 |
| c.935C>G<br>p.(Thr312Ser) | 141102 | Conflicting | LB/VUS | B/LB | B/LB | B | BS1, BS3, BP4 |
| c.892G>A<br>p.(Glu298Lys) | 141483 | Conflicting | LB | LB | LB | LB | BP4, BS3 |
| c.704A>G<br>p.(Asn235Ser) | 127821 | Conflicting | LB/VUS | VUS | B | B | BS1, BS3, BS4,<br>BP4, BP2 |
| c.245C>T<br>p.(Pro82Leu) | 182946 | Conflicting | LB/VUS | VUS | LB | LB | BS3, BP4 |
| c.28G>A<br>p.(Val10Ile) | 127806 | Conflicting | LB/VUS | LB | LB/VUS | LB | BS3, BP4 |
| c.21T>A<br>p.(Asp7Glu) | 140782 | Conflicting | LB | VUS | VUS | LB | PM2_Supporting,<br>BS3, BP4 |
| c.145G>A<br>p.(Asp49Asn) | 186363 | Conflicting | LB | VUS | LB | LB | BP4, BS3 |
| c.641A>G<br>p.(His214Arg) | 376615 | Conflicting | LP | LP/P | LP | LP | PS3,<br>PM2_Supporting,<br>PS4_Supporting |
| c.217G>A<br>p.(Val73Met) | 142386 | Conflicting | LB | VUS | LB/VUS | B | BS1, BS3, BP4 |
| c.139C>T<br>p.(Pro47Ser) | 43588 | Conflicting | B | B/LB | B | B | BA1, BS3, BP4 |
| c.319T>C<br>p.(Tyr107His) | 140786 | Conflicting | LB/VUS | LB/VUS | B | B | BA1, BP4,<br>BS3_Supporting |
| c.428T>C<br>p.(Val143Ala) | 792574 | NA | NA | LP/P | LP | LP | PS3, PM6,<br>PM2_Supporting,<br>PP3 |

Abbreviations: B = Benign, LB = Likely benign, VUS = Variant of unknown significance, LP = Likely pathogenic, P = Pathogenic, Conflicting = Conflicting interpretations of pathogenicity, NA = Not Available.
\*ClinVar assertions were documented in September 2019. Conflicting variants were those that spanned different clinically relevant classifications.
^Assertions provided by VCEP members with variant nominations. Some variants were nominated by multiple members; some with conflicting assertions. Variants listed as “NA” were not nominated by a VCEP member.
#Variants were assessed by two curators using the original ACMG/AMP guidelines and any curation assertions conflicts are noted.

#### Cis/trans testing data

The use of BP2, which uses observation with another pathogenic variant as evidence against pathogenicity, was specified as follows: variant is observed *in trans* with a *TP53* pathogenic variant (phase confirmed), or there are three or more observations with a *TP53* pathogenic variant when phase is unknown. In this scenario, the variant must be seen with at least two different *TP53* pathogenic variants (as specified by the *TP53* VCEP).

### Testing of the *TP53* specifications on a pilot set of variants

There were 43 variants used for pilot testing. Classifications were compared between biocurators using the original ACMG/AMP and the *TP53-*specific guidelines, the existing assertions in ClinVar, and prior assertions by experts (Table 4).

Intra-biocurator consistency of variant classification was high: 72.1% using the original ACMG/AMP guidelines and 81.4% using *TP53*-specific guidelines (Table 4). There were fewer VUS using *TP53*-specific guidelines compared with the original guidelines (5/43 (12%) vs 12/43 (28%)) (Figure 2). Of the 42 pilot *TP53* variants recorded in ClinVar in September 2019, there were 16 (38%) clinically relevant discrepancies in ClinVar at that time (see Table 4). Using the *TP53*-specific guidelines and sharing clinical data amongst experts and clinical laboratories, the majority of VUS were downgraded to LB (13/16; 81%), two moved to LP (2/16; 12.5%), and one remained at VUS (1/16; 6%).

**Figure 1.**
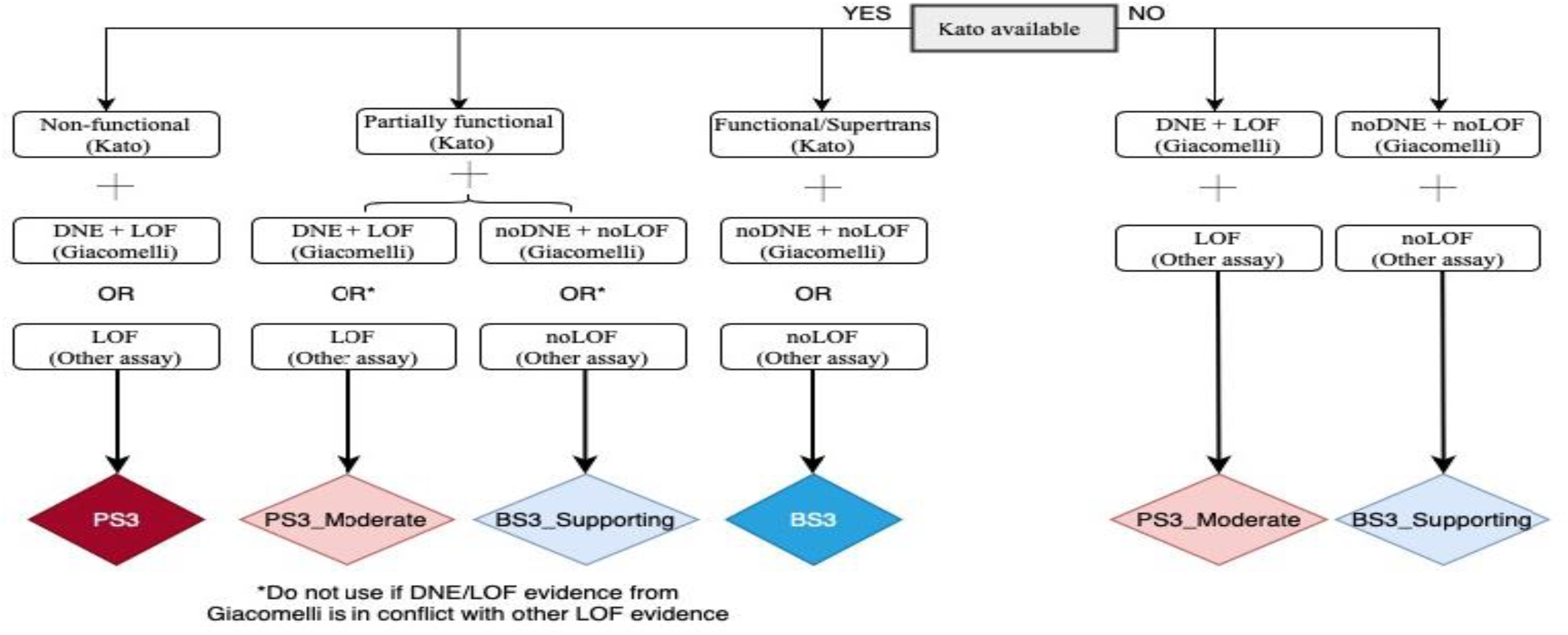
Flow chart for the specifications of PS3 and BS3 criteria. Non-functional (Kato) = median transactivation activity ≤20%; Partially functional (Kato) = median transactivation activity >20 and ≤75%; Functional/Supertrans (Kato) = median transactivation activity >75%; DNE+LOF (Giacomelli) = p53WTNutlin3 Z-score ≥ 0.61 and Etoposide Z-score ≤ -0.21 = noDNE+noLOF (Giacomelli) = p53WTNutlin3 Z-score < 0.61 and Etoposide Z-score >-0.21. Other assays are available in IARC *TP53* Database or original publications, and include *in vitro* growth assays in H1299 human cells from Kotler et al., (2018) with RFS score ≥ -1.0 for LOF and RFS score < -1.0 for noLOF; or colony formation assays, growth suppression assays, apoptosis assays, tetramer assays, knock-in mouse models.*If a variant does not match any of the possibilities shown, it is considered to have “no evidence to review” and no functional criterion can be applied. Abbreviations: DNE = Dominant-negative effect; LOF = Loss-of-function.

**Figure 2.**
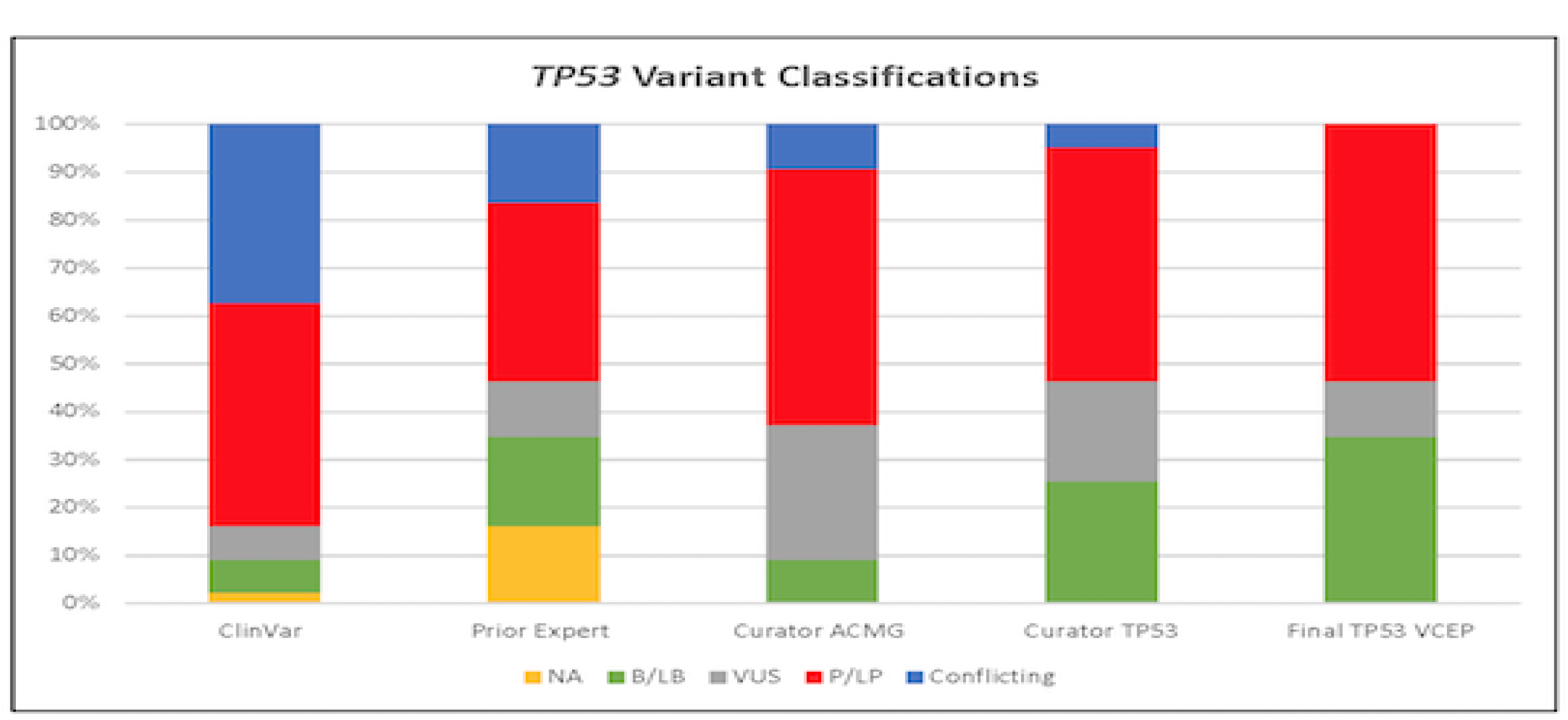
Variant classifications for 43 pilot *TP53* variants in ClinVar, from the nominating expert(s), biocurators using the original ACMG/AMP guidelines, and the *TP53*-specific guidelines. Abbreviations: B = Benign, LB = Likely benign, VUS = Variant of unknown significance, LP = Likely pathogenic, P = Pathogenic, Conflicting = Clinically relevant conflicting interpretations of pathogenicity, and NA = Not Available.

## DISCUSSION

The task of the *TP53* VCEP was to specify the ACMG/AMP guidelines to assist with the clinical classification of variants in the *TP53* gene. There is a rapidly growing number of individuals without personal or family history consistent with LFS who have germline *TP53* variants (Batalini et al., 2019; Fortuno, James, & Spurdle, 2018), and cancer surveillance for people with *TP53* pathogenic germline variants is time intensive and emotionally stressful (Ballinger et al., 2017; Kratz et al., 2017; Pantaleao et al., 2019; Villani et al., 2016; Young et al., 2019). Therefore, robust *TP53* variant classification is essential for optimal clinical care.

Our pilot study demonstrated that the *TP53*-specific guidelines decreased the number of VUS compared with the original guidelines and increased the number of variants classified as not clinically relevant. Our study also showed strong intra-biocurator consistency, suggesting that the criteria appear to be straightforward to interpret; this should allow for fewer conflicting variant calls between laboratories.

One of the benefits of curating *TP53* was the wealth of existing knowledge including, data readily available in public databases. The IARC *TP53* database is a rich source of data that was helpful in assessing pathogenicity codes, including PS1, PS2, PS4, PM5, PM6, and PP1. The tremendous amount of data on p53 mouse models searchable by knockout variant of interest was also useful for adapting functional rule codes. Additionally, the *TP53* VCEP benefited from data sharing amongst participating researchers, clinicians, and clinical laboratories. VCEP members added information pertaining to additional probands to those identified in the literature, to increase the number of pathogenic codes applicable to certain variants and to shift classifications from VUS to LP. Laboratory members also shared data from their large hereditary cancer panels that were helpful in using the benign codes BS2 and BP2.

Consistent with the ClinGen VCEP process, all of the *TP53* classified variants are present in the ClinVar database with a summary of the classification process. More details about the evidence used to classify the variant is available through a link to the public access ClinGen Evidence Repository (https://erepo.clinicalgenome.org/evrepo/). The *TP53* VCEP will continue to meet monthly to curate additional variants and submit them to ClinVar. Variants classified as LP or VUS will be reviewed at least every two years in the event new evidence has emerged, while variants classified as LB will be reviewed when new evidence is available or when requested by the public via the ClinGen website. It is also anticipated that these specifications will be reviewed as needed, to incorporate additional considerations and evidence types to improve *TP53* variant classification (Fortuno, Cipponi, et al., 2019; Fortuno et al., 2020). The most up to date version of the ClinGen *TP53* VCEP specifications is available at https://clinicalgenome.org/affiliation/50013/.

## Data Availability

Most of the data that support the findings of this study are openly available in the IARC TP53 Database at https://p53.iarc.fr (version R20, July 2019). Other data supporting the findings of this study are not publicly available due to private or ethical restrictions. The publicly available Clinical Genome Resource (ClinGen) Evidence Repository contains all of the curated evidence for the variants submitted to ClinVar (e.g., https://erepo.clinicalgenome.org/evrepo/ui/interpretation/7b4332f9-03a6-43f1-afde-508d91bd92d5).

## FUNDING

This Expert Panel is funded by National Human Genome Research & National Cancer Institutes (1U41HG006834, 1U01HG007437, 1U01HG007436, HHSN261200800001E, U41HG009650). The work of C.F. was supported by a University of Queensland (UQ) International Scholarship from the UQ School of Medicine. The work of M.F., K.C.A., and S.A.S. was supported by the intramural research program of the Division of Cancer Epidemiology and Genetics, National Cancer Institute, National Institutes of Health. The work of A.B.S. was supported by Australian National Health and Medical Research funding (ID1061779, ID1161589). The work of D.G.E. was supported by the NIHR Manchester Biomedical Research Centre (IS-BRC-1215–20007). The work of T.P.S. was supported by NIH-NCI K08CA234394 and R01CA242218.

## ACKNOWLEDGEMENTS

The ClinGen *TP53* Variant Curation Expert Panel would like to thank the ClinGen *PTEN* Variant Curation Expert Panel and the Executive Committee of the Hereditary Cancer Clinical Domain Working Group for sharing their experience with developing criteria specifications. We would also like to thank Steven Harrison, Leslie Biesecker and the remainder of the ClinGen Sequence Variant Interpretation Working Group for their guidance. Finally, we wish to thank present and past members of the *TP53* VCEP, Maria Isabel Achatz, Rebecca Bassett, Jeffrey Bissonnette, David Goldgar, Sharisse Jimenez, Chimene Kesserwan, Deb Ritter, Leighton Telling and Mackenzie Trapp.

## CONFLICTS OF INTEREST

The following authors have no conflicts of interest to disclose: K.L., M.O., K.C.A., S.B., A.K.M.F., M.F., B.A.S., L.W., L.Z. The following authors have made extensive contributions to the *TP53* literature and have previously published assertions on *TP53* variants: C.F., P.L.M., L.D.A., D.G.E., P.J., K.M., T.P.S., A.B.S., S.A.S. The following authors are an employee, trainee or consultant for a commercial laboratory that offers genetic testing for *TP53*: T.P., R.H., K.M., S.E.P. J.M. is an employee of GeneDx/BioReference Laboratories, Inc./OPKO Health and has a salary as the only disclosure. The PERCH software, for which B.J.F. is the inventor, has been non-exclusively licensed to Ambry Genetics Corporation for their clinical genetic testing services and research. B.J.F. also reports funding and sponsorship to his institution on his behalf from Pfizer Inc. and Regeneron Genetics Center LLC.

## DATA AVAILABILITY STATEMENT

Most of the data that support the findings of this study are openly available in the IARC TP53 Database at https://p53.iarc.fr (version R20, July 2019). Other data supporting the findings of this study are not publicly available due to private or ethical restrictions. The publicly available Clinical Genome Resource (ClinGen) Evidence Repository contains all of the curated evidence for the variants submitted to ClinVar (*e*.*g*., *https://erepo.clinicalgenome.org/evrepo/ui/interpretation/7b4332f9-03a6-43f1-afde-508d91bd92d5)*.

## REFERENCES

Abou Tayoun, A. N., Pesaran, T., DiStefano, M. T., Oza, A., Rehm, H. L., Biesecker, L. G., & Harrison, S. M. (2018). Recommendations for interpreting the loss of function PVS1 ACMG/AMP variant criterion. Hum Mutat, 39(11), 1517–1524. doi:10.1002/humu.23626

Amendola, L. M., Dorschner, M. O., Robertson, P. D., Salama, J. S., Hart, R., Shirts, B. H., … Jarvik, G. P. (2015). Actionable exomic incidental findings in 6503 participants: challenges of variant classification. Genome Res, 25(3), 305–315. doi:10.1101/gr.183483.114

Ballinger, M. L., Best, A., Mai, P. L., Khincha, P. P., Loud, J. T., Peters, J. A., … Savage, S. A. (2017). Baseline Surveillance in Li-Fraumeni Syndrome Using Whole-Body Magnetic Resonance Imaging: A Meta-analysis. JAMA Oncol. doi:10.1001/jamaoncol.2017.1968

Batalini, F., Peacock, E. G., Stobie, L., Robertson, A., Garber, J., Weitzel, J. N., & Tung, N. M. (2019). Li-Fraumeni syndrome: not a straightforward diagnosis anymore-the interpretation of pathogenic variants of low allele frequency and the differences between germline PVs, mosaicism, and clonal hematopoiesis. Breast Cancer Res, 21(1), 107. doi:10.1186/s13058-019-1193-1

Biesecker, L. G., & Harrison, S. M. (2018). The ACMG/AMP reputable source criteria for the interpretation of sequence variants. Genet Med, 20(12), 1687–1688. doi:10.1038/gim.2018.42

Bittar, C. M., Vieira, I. A., Sabato, C. S., Andreis, T. F., Alemar, B., Artigalas, O., … Ashton-Prolla, P. (2019). TP53 variants of uncertain significance: increasing challenges in variant interpretation and genetic counseling. Fam Cancer, 18(4), 451–456. doi:10.1007/s10689-019-00140-w

Bouaoun, L., Sonkin, D., Ardin, M., Hollstein, M., Byrnes, G., Zavadil, J., & Olivier, M. (2016). TP53 Variations in Human Cancers: New Lessons from the IARC TP53 Database and Genomics Data. Hum Mutat, 37(9), 865–876. doi:10.1002/humu.23035

Bougeard, G., Renaux-Petel, M., Flaman, J. M., Charbonnier, C., Fermey, P., Belotti, M., … Frebourg, T. (2015). Revisiting Li-Fraumeni Syndrome From TP53 Mutation Carriers. J Clin Oncol, 33(21), 2345–2352. doi:10.1200/JCO.2014.59.5728

Brosh, R., & Rotter, V. (2009). When mutants gain new powers: news from the mutant p53 field. Nat Rev Cancer, 9(10), 701–713. doi:10.1038/nrc2693

Caron, O., Frébourg, T., Benusiglio, P. R., Faivre, L., Dugast, C., Bonadona, V., … Brugieres, L. (2016). The 2016 Li-Fraumeni syndrome cancer spectrum. Journal of Clinical Oncology, 34(15_suppl), 1535–1535. doi:10.1200/JCO.2016.34.15_suppl.1535

Chang, M. T., Asthana, S., Gao, S. P., Lee, B. H., Chapman, J. S., Kandoth, C., … Taylor, B. S. (2016). Identifying recurrent mutations in cancer reveals widespread lineage diversity and mutational specificity. Nat Biotechnol, 34(2), 155–163. doi:10.1038/nbt.3391

Chang, M. T., Bhattarai, T. S., Schram, A. M., Bielski, C. M., Donoghue, M. T. A., Jonsson, P., … Taylor, B. S. (2018). Accelerating Discovery of Functional Mutant Alleles in Cancer. Cancer Discov, 8(2), 174–183. doi:10.1158/2159-8290.CD-17-0321

Coffee, B., Cox, H. C., Kidd, J., Sizemore, S., Brown, K., Manley, S., & Mancini-DiNardo, D. (2017). Detection of somatic variants in peripheral blood lymphocytes using a next generation sequencing multigene pan cancer panel. Cancer Genet, 211, 5–8. doi:10.1016/j.cancergen.2017.01.002

de Andrade, K. C., Frone, M. N., Wegman-Ostrosky, T., Khincha, P. P., Kim, J., Amadou, A., … Achatz, M. I. (2019). Variable population prevalence estimates of germline TP53 variants: A gnomAD-based analysis. Hum Mutat, 40(1), 97–105. doi:10.1002/humu.23673

de Andrade, K. C., Mirabello, L., Stewart, D. R., Karlins, E., Koster, R., Wang, M., … Achatz, M. I. (2017). Higher-than-expected population prevalence of potentially pathogenic germline TP53 variants in individuals unselected for cancer history. Hum Mutat, 38(12), 1723–1730. doi:10.1002/humu.23320

Desmet, F. O., Hamroun, D., Lalande, M., Collod-Beroud, G., Claustres, M., & Beroud, C. (2009). Human Splicing Finder: an online bioinformatics tool to predict splicing signals. Nucleic Acids Res, 37(9), e67. doi:10.1093/nar/gkp215

Evans, D. G., Birch, J. M., Ramsden, R. T., Sharif, S., & Baser, M. E. (2006). Malignant transformation and new primary tumours after therapeutic radiation for benign disease: substantial risks in certain tumour prone syndromes. J Med Genet, 43(4), 289–294. doi:10.1136/jmg.2005.036319

Feng, B. J. (2017). PERCH: A Unified Framework for Disease Gene Prioritization. Hum Mutat, 38(3), 243–251. doi:10.1002/humu.23158

Fortuno, C., Cipponi, A., Ballinger, M. L., Tavtigian, S. V., Olivier, M., Ruparel, V., … James, P. A. (2019). A quantitative model to predict pathogenicity of missense variants in the TP53 gene. Hum Mutat, 40(6), 788–800. doi:10.1002/humu.23739

Fortuno, C., James, P. A., & Spurdle, A. B. (2018). Current review of TP53 pathogenic germline variants in breast cancer patients outside Li-Fraumeni syndrome. Hum Mutat, 39(12), 1764–1773. doi:10.1002/humu.23656

Fortuno, C., James, P. A., Young, E. L., Feng, B., Olivier, M., Pesaran, T., … Spurdle, A. B. (2018). Improved, ACMG-Compliant, in silico prediction of pathogenicity for missense substitutions encoded by TP53 variants. Hum Mutat. doi:10.1002/humu.23553

Fortuno, C., Pesaran, T., Dolinsky, J., Yussuf, A., McGoldrick, K., Goldgar, D., … Spurdle, B. (2020). Differences in patient ascertainment affect the use of gene-specified ACMG/AMP phenotype-related variant classification criteria: evidence for TP53. Hum Mutat. doi:10.1002/humu.23972

Fortuno, C., Pesaran, T., Dolinsky, J., Yussuf, A., McGoldrick, K., Kho, P. F., … Spurdle, A. (2019). p53 major hotspot variants are associated with poorer prognostic features in hereditary cancer patients. Cancer Genet, 235–236, 21-27. doi:10.1016/j.cancergen.2019.05.002

Freed-Pastor, W. A., & Prives, C. (2012). Mutant p53: one name, many proteins. Genes Dev, 26(12), 1268–1286. doi:10.1101/gad.190678.112

Giacomelli, A. O., Yang, X., Lintner, R. E., McFarland, J. M., Duby, M., Kim, J., Hahn, W. C. (2018). Mutational processes shape the landscape of TP53 mutations in human cancer. Nat Genet, 50(10), 1381–1387. doi:10.1038/s41588-018-0204-y

Gonzalez, K. D., Buzin, C. H., Noltner, K. A., Gu, D., Li, W., Malkin, D., & Sommer, S. S. (2009). High frequency of de novo mutations in Li-Fraumeni syndrome. J Med Genet, 46(10), 689–693. doi:10.1136/jmg.2008.058958

Gonzalez, K. D., Noltner, K. A., Buzin, C. H., Gu, D., Wen-Fong, C. Y., Nguyen, V. Q., … Weitzel, J. N. (2009). Beyond Li Fraumeni Syndrome: clinical characteristics of families with p53 germline mutations. J Clin Oncol, 27(8), 1250–1256. doi:10.1200/JCO.2008.16.6959

Grantham, R. (1974). Amino acid difference formula to help explain protein evolution. Science, 185(4154), 862–864. doi:10.1126/science.185.4154.862

Kaariainen, H., Muilu, J., Perola, M., & Kristiansson, K. (2017). Genetics in an isolated population like Finland: a different basis for genomic medicine? J Community Genet, 8(4), 319–326. doi:10.1007/s12687-017-0318-4

Kalia, S. S., Adelman, K., Bale, S. J., Chung, W. K., Eng, C., Evans, J. P., … Miller, D. T. (2017). Recommendations for reporting of secondary findings in clinical exome and genome sequencing, 2016 update (ACMG SF v2.0): a policy statement of the American College of Medical Genetics and Genomics. Genet Med, 19(2), 249–255. doi:10.1038/gim.2016.190

Karczewski, K. J., Francioli, L. C., Tiao, G., Cummings, B. B., Alföldi, J., Wang, Q., … MacArthur, D. G. (2019). Variation across 141,456 human exomes and genomes reveals the spectrum of loss-of-function intolerance across human protein-coding genes. bioRxiv, 531210. doi:10.1101/531210

Kato, S., Han, S. Y., Liu, W., Otsuka, K., Shibata, H., Kanamaru, R., & Ishioka, C. (2003). Understanding the function-structure and function-mutation relationships of p53 tumor suppressor protein by high-resolution missense mutation analysis. Proc Natl Acad Sci U S A, 100(14), 8424–8429. doi:10.1073/pnas.1431692100

Kotler, E., Shani, O., Goldfeld, G., Lotan-Pompan, M., Tarcic, O., Gershoni, A., … Segal, E. (2018). A Systematic p53 Mutation Library Links Differential Functional Impact to Cancer Mutation Pattern and Evolutionary Conservation. Mol Cell, 71(1), 178–190 e178. doi:10.1016/j.molcel.2018.06.012

Kratz, C. P., Achatz, M. I., Brugieres, L., Frebourg, T., Garber, J. E., Greer, M. C., … Malkin, D. (2017). Cancer Screening Recommendations for Individuals with Li-Fraumeni Syndrome. Clin Cancer Res, 23(11), e38–e45. doi:10.1158/1078-0432.ccr-17-0408

Lalloo, F., Varley, J., Moran, A., Ellis, D., O’Dair, L., Pharoah, P., … Evans, D. G. (2006). BRCA1, BRCA2 and TP53 mutations in very early-onset breast cancer with associated risks to relatives. Eur J Cancer, 42(8), 1143–1150. doi:10.1016/j.ejca.2005.11.032

Landrum, M. J., Lee, J. M., Benson, M., Brown, G. R., Chao, C., Chitipiralla, S., … Maglott, D. R. (2018). ClinVar: improving access to variant interpretations and supporting evidence. Nucleic Acids Res, 46(D1), D1062–D1067. doi:10.1093/nar/gkx1153

Lane, D. P. (1992). Cancer. p53, guardian of the genome. Nature, 358(6381), 15–16. doi:10.1038/358015a0

Lee, K., Krempely, K., Roberts, M. E., Anderson, M. J., Carneiro, F., Chao, E., … Karam, R. (2018). Specifications of the ACMG/AMP variant curation guidelines for the analysis of germline CDH1 sequence variants. Hum Mutat, 39(11), 1553–1568. doi:10.1002/humu.23650

Li, F. P., Fraumeni, J. F., Jr., Mulvihill, J. J., Blattner, W. A., Dreyfus, M. G., Tucker, M. A., & Miller, R. W. (1988). A cancer family syndrome in twenty-four kindreds. Cancer Res, 48(18), 5358-5362. Retrieved from http://www.ncbi.nlm.nih.gov/pubmed/3409256

Mai, P. L., Best, A. F., Peters, J. A., DeCastro, R. M., Khincha, P. P., Loud, J. T., … Savage, S. A. (2016). Risks of first and subsequent cancers among TP53 mutation carriers in the National Cancer Institute Li-Fraumeni syndrome cohort. Cancer, 122(23), 3673–3681. doi:10.1002/cncr.30248

Mathe, E., Olivier, M., Kato, S., Ishioka, C., Hainaut, P., & Tavtigian, S. V. (2006). Computational approaches for predicting the biological effect of p53 missense mutations: a comparison of three sequence analysis based methods. Nucleic Acids Res, 34(5), 1317–1325. doi:10.1093/nar/gkj518

Mester, J. L., Ghosh, R., Pesaran, T., Huether, R., Karam, R., Hruska, K. S., … Eng, C. (2018). Gene-specific criteria for PTEN variant curation: Recommendations from the ClinGen PTEN Expert Panel. Hum Mutat, 39(11), 1581–1592. doi:10.1002/humu.23636

Olivier, M., Hollstein, M., & Hainaut, P. (2010). TP53 mutations in human cancers: origins, consequences, and clinical use. Cold Spring Harb Perspect Biol, 2(1), a001008. doi:10.1101/cshperspect.a001008

Pantaleao, A., Young, J. L., Epstein, N. B., Carlson, M., Bremer, R. C., Khincha, P. P., … Werner-Lin, A. (2019). Family Health Leaders: Lessons on Living with Li-Fraumeni Syndrome across Generations. Fam Process. doi:10.1111/famp.12497

Richards, S., Aziz, N., Bale, S., Bick, D., Das, S., Gastier-Foster, J., … Committee, A. L. Q. A. (2015). Standards and guidelines for the interpretation of sequence variants: a joint consensus recommendation of the American College of Medical Genetics and Genomics and the Association for Molecular Pathology. Genet Med, 17(5), 405–424. doi:10.1038/gim.2015.30

Schneider, K., Zelley, K., Nichols, K. E., & Garber, J. (1993). Li-Fraumeni Syndrome. In R. A. Pagon, M. P. Adam, H. H. Ardinger, S. E. Wallace, A. Amemiya, L. J. H. Bean, T. D. Bird, C. T. Fong, H. C. Mefford, R. J. H. Smith, & K. Stephens (Eds.), GeneReviews(R). Seattle (WA).

Schon, K., & Tischkowitz, M. (2018). Clinical implications of germline mutations in breast cancer: TP53. Breast Cancer Res Treat, 167(2), 417–423. doi:10.1007/s10549-017-4531-y

Shamsani, J., Kazakoff, S. H., Armean, I. M., McLaren, W., Parsons, M. T., Thompson, B. A., … Spurdle, A. B. (2019). A plugin for the Ensembl Variant Effect Predictor that uses MaxEntScan to predict variant spliceogenicity. Bioinformatics, 35(13), 2315–2317. doi:10.1093/bioinformatics/bty960

Shi, L., Webb, B. D., Birch, A. H., Elkhoury, L., McCarthy, J., Cai, X., … Kornreich, R. (2017). Comprehensive population screening in the Ashkenazi Jewish population for recurrent disease-causing variants. Clin Genet, 91(4), 599–604. doi:10.1111/cge.12834

Tavtigian, S. V., Greenblatt, M. S., Harrison, S. M., Nussbaum, R. L., Prabhu, S. A., Boucher, K. M., & Biesecker, L. G. (2018). Modeling the ACMG/AMP variant classification guidelines as a Bayesian classification framework. Genet Med. doi:10.1038/gim.2017.210

Villani, A., Shore, A., Wasserman, J. D., Stephens, D., Kim, R. H., Druker, H., … Malkin, D. (2016). Biochemical and imaging surveillance in germline TP53 mutation carriers with Li-Fraumeni syndrome: 11 year follow-up of a prospective observational study. Lancet Oncol, 17(9), 1295–1305. doi:10.1016/S1470-2045(16)30249-2

Walsh, M. F., Ritter, D. I., Kesserwan, C., Sonkin, D., Chakravarty, D., Chao, E., … Plon, S. E. (2018). Integrating somatic variant data and biomarkers for germline variant classification in cancer predisposition genes. Hum Mutat, 39(11), 1542–1552. doi:10.1002/humu.23640

Weitzel, J. N., Chao, E. C., Nehoray, B., Van Tongeren, L. R., LaDuca, H., Blazer, K. R., … Jasperson, K. (2017). Somatic TP53 variants frequently confound germ-line testing results. Genet Med. doi:10.1038/gim.2017.196

Whiffin, N., Minikel, E., Walsh, R., O’Donnell-Luria, A. H., Karczewski, K., Ing, A. Y., … Ware, J. S. (2017). Using high-resolution variant frequencies to empower clinical genome interpretation. Genet Med. doi:10.1038/gim.2017.26

Yeo, G., & Burge, C. B. (2004). Maximum entropy modeling of short sequence motifs with applications to RNA splicing signals. J Comput Biol, 11(2-3), 377–394. doi:10.1089/1066527041410418

Young, J. L., Pantaleao, A., Zaspel, L., Bayer, J., Peters, J. A., Khincha, P. P., … Werner-Lin, A. (2019). Couples coping with screening burden and diagnostic uncertainty in Li-Fraumeni syndrome: Connection versus independence. J Psychosoc Oncol, 37(2), 178–193. doi:10.1080/07347332.2018.1543376

Zerdoumi, Y., Lanos, R., Raad, S., Flaman, J. M., Bougeard, G., Frebourg, T., & Tournier, I. (2017). Germline TP53 mutations result into a constitutive defect of p53 DNA binding and transcriptional response to DNA damage. Hum Mol Genet, 26(14), 2591–2602. doi:10.1093/hmg/ddx106

